# COVID-19 versus non-COVID-19 pneumonia: A retrospective cohort study in Chengdu, China

**DOI:** 10.1101/2020.04.28.20082784

**Authors:** Xiao-Jin Li, Bing-Xing Shuai, Zhong-Wei Zhang, Yan Kang

## Abstract

**Background and Objective:** Since the outbreak of coronavirus disease 2019 (COVID-19) in December 2019 in Wuhan, Hubei Province, China, it has spread throughout the world and become a global public health emergency. It is important to distinguish COVID-19 from other viral pneumonias to properly screen and diagnose patients, reduce nosocomial infections, and complement the inadequacy of nucleic acid testing. In this study, we retrospectively analysed the clinical data of COVID-19 versus non-COVID-19 patients treated at our hospital between January 17 and February 27, 2020 to summarize our clinical experience in the differential diagnosis of COVID-19.

**Methods:** In this retrospective cohort study, 23 confirmed COVID-19 patients were consecutively enrolled from January 17 to February 27, 2020, and 29 confirmed non-COVID-19 patients were enrolled in the West China Hospital of Sichuan University. We collected baseline data, epidemiological data, clinical characteristics, imaging findings, viral nucleic acid test results, and survival data. SPSS v22.0 was used for the statistical analysis. Outcomes were followed-up until March 25.

**Results:** A total of 52 patients were included in this study, including 23 COVID-19 patients and 29 non-COVID-19 patients. No significant between-group differences were observed for age, sex, primary signs or symptoms, cellular immunity, or platelet count. Significant between-group differences were observed in clinical characteristics such as dry cough, contact with individuals from Wuhan, some underlying diseases, nucleated cell count, chest imaging findings, viral nucleic acid test results, 28-day mortality, and 28-day survival.

**Conclusion:** Epidemiological data, clinical symptoms, nucleic acid test results for COVID-19 and chest CT manifestation may help distinguish COVID-19 from non-COVID-19 cases, prevent imported cases and nosocomial infections.

## Introduction

Coronavirus disease 2019 (COVID-19) was first detected in December 2019 in Wuhan, Hubei Province, China[1]. The virus spread to other regions of China via human-to-human transmission[2, 3], resulting in imported cases and clusters of cases. Since then, COVID-19 has spread rapidly throughout the world. As of March 31, 700,000 confirmed cases had been reported worldwide. The outbreak occurred along with seasonal infections such as influenza and other transmissible viral respiratory diseases[4]. Therefore, it is important to distinguish different types of viral pneumonia. Given the rapid spread of COVID-19 and its significant impact on public health[5, 6], some researchers have collected data from different hospitals and investigated the clinical characteristics of COVID-19 versus bacterial pneumonia[7]. However, the researchers did not compare COVID-19 and other viral pneumonia cases that occurred during the same period. COVID-19 is highly contagious[6, 8, 9] and spreads quickly[10, 11] without apparent symptoms in some cases[12, 13]. Nucleic acid testing is currently the main method of diagnosis[14]. Early nucleic acid testing plays a key role in detecting patients early and facilitating their early quarantine and treatment[15]. However, nucleic acid testing has some limitations, including a high false-negative rate[16] and variations in the quality of reagents from different sources. Epidemiological data, chest imaging findings, and clinical characteristics must be taken into consideration to make a diagnosis. It is important to distinguish COVID-19 from other types of viral pneumonia to properly screen and diagnose patients and reduce nosocomial infections. In this study, we retrospectively analysed the clinical data of COVID-19 versus non-COVID-19 patients treated at our hospital between January 17 and February 27, 2020 to provide a reference for the differential diagnosis of COVID-19, help evaluate the prevention and control of nosocomial infections at our hospital, and add to the published clinical experience.

## Methods

### Ethics review

This retrospective study was approved by the Ethics Committee of the West China Hospital, Sichuan University, on February 10, 2020 (EC Number: 2019 No. 130).

### Data collection

We searched electronic medical records in the medical information database of our hospital with the following keywords: unexplained fever, severe acute respiratory syndrome, coronavirus 2 (SARS-CoV-2), viral pneumonia, respiratory infection, bronchitis, pulmonary infection, pneumonia, severe pneumonia, and acute respiratory distress syndrome (ARDS). We retrieved and retrospectively analysed case data (per patient ID), including the baseline data, epidemiological data, clinical characteristics, laboratory test results, imaging findings, and results of nucleic acid tests for SARS-CoV-2. The data were entered in a standardized data collection form and then independently reviewed by two investigators to check the data accuracy and the availability of epidemiological data and information about clinical symptoms. In case of missing data, the investigators contacted the attending physicians and patients or their families directly to obtain accurate data.

### Sample collection for virus tests and chest computed tomography (CT)

A throat swab or bronchoalveolar lavage fluid was collected according to the standard sampling requirements and procedures from all patients admitted to the West China Hospital, Sichuan University, between January 17 and February 27, 2020. The samples were sealed and sent to the clinical laboratory for the screening of 11 viruses (influenza A virus, adenovirus, bocavirus, rhinovirus, influenza A virus subtype H1N1, parainfluenza virus, metapneumovirus, influenza B virus, influenza A virus subtype H3N2, coronavirus, respiratory syncytial virus) and for the nucleic acid test for SARS-CoV-2. Moreover, all patients underwent chest CT.

The inclusion and exclusion criteria are shown in the study protocol (Figure 1). Baseline data were collected, including age, sex, underlying diseases, and history of travelling to or from Wuhan or contact with individuals from Hubei. Clinical symptoms included cough, fever, difficulty breathing, chest pain, chest tightness, headache, muscle aches, fatigue, and diarrhoea. Laboratory tests included white blood cell count (WBC), neutrophil count and percentage, lymphocyte count, cellular immunity (CD4:CD8 ratio), platelet count, procalcitonin (PCT), alanine aminotransferase (ALT), lactate dehydrogenase (LDH), and creatinine (Cr). Moreover, chest imaging findings, viral nucleic acid test results, 28-day mortality, and 28-day survival were collected. We analysed and compared these data in COVID-19 versus non-COVID-19 patients.

**Figure 1.**
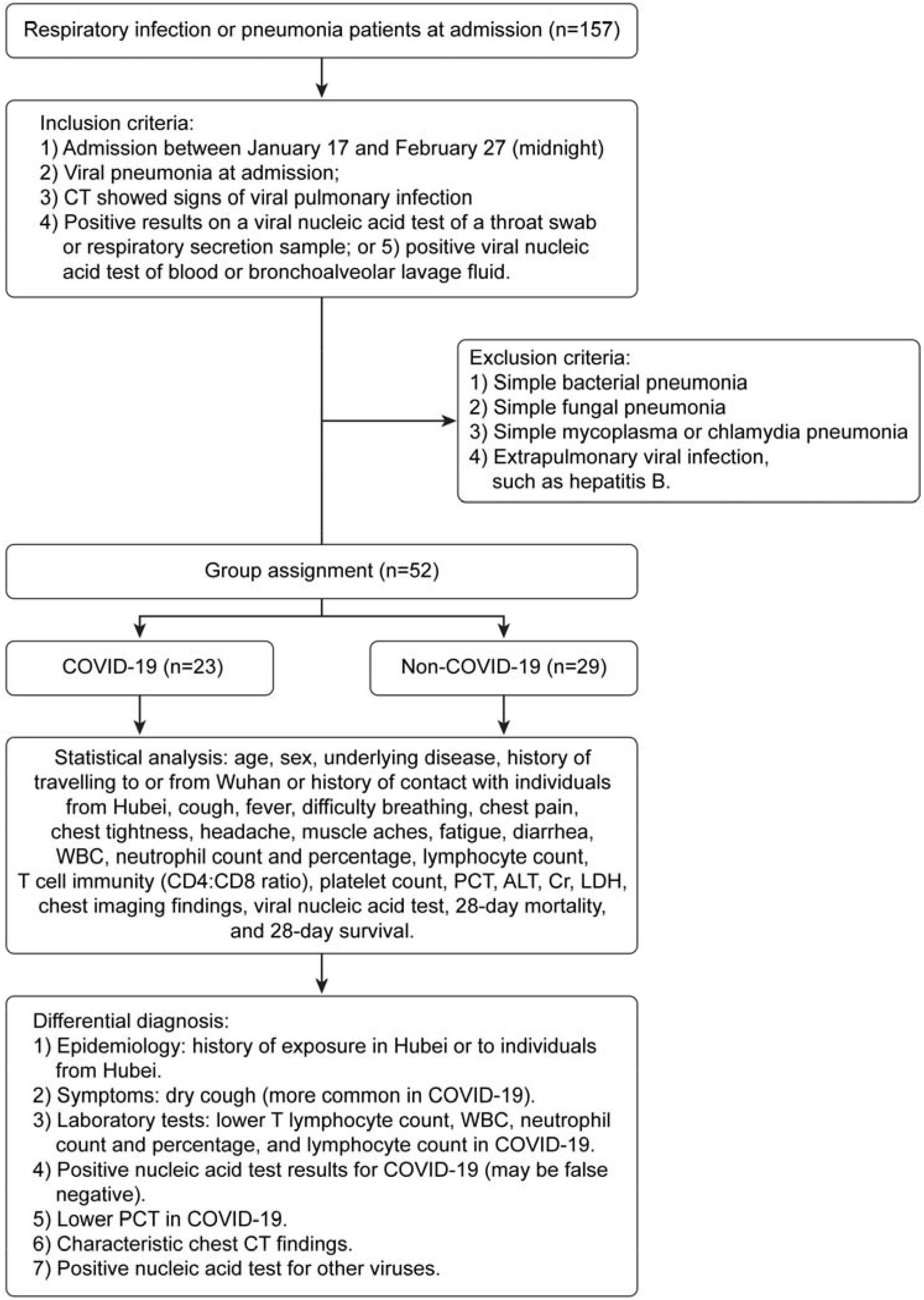
Flow chart of study enrolment

### Statistical analysis

Pearson’s chi-squared test or Fisher’s exact probability method was used to compare count data between groups. Normally distributed data are expressed as the mean ± standard deviation. A one-way ANOVA or Student’s t-test was used for measurement data. Non-normal data were compared using the Wilcoxon rank-sum test. The 28-day mortality test was conducted using a log-rank Kaplan-Meier analysis of the logarithmic survival curve. *P* values less than 0.05 indicated statistical significance. The statistical tool used was SPSS 22.0 (SPSS Science Inc., Chicago, IL, USA).

### Results

We searched the medical information database of our hospital with relevant keywords and retrieved 157 cases treated at our hospital between January 17 and February 27, 2020. Among them, 52 patients were diagnosed with viral pneumonia based on positive nucleic acid test results and chest CT and thus met the inclusion and exclusion criteria of this study. Specifically, 23 patients (44.23%) were diagnosed with COVID-19. Among them, one patient also had a parainfluenza virus infection, and one patient also had an influenza A/B virus infection. All COVID-19 patients were admitted to the isolation ward of the Department of Infectious Diseases. During the same period, 29 patients (55.77%) diagnosed with other viral pneumonias (non-COVID-19) were admitted to our hospital. Among them, 12 patients had cytomegalovirus infection, eight had influenza A (H3N2) infection, four had rhinovirus infection, four had EpsteinͰBarr virus infection, three had respiratory syncytial virus infection, one patient had influenza B infection, one patient had adenovirus infection, and one patient had metapneumovirus infection(Table 2). All of these cases are common viral respiratory infections in the winter.

No significant between-group differences were observed in age, sex, fever, muscle aches, fatigue, headache, chest pain, diarrhoea, heart rate, blood pressure, respiratory rate, cellular immunity, platelet count, ALT, or liver or kidney function (Table 1). Significant between-group differences were observed in dry cough, difficulty breathing, history of travelling to or from Wuhan, history of contact with individuals from Hubei, some underlying diseases, viral nucleic acid test result, WBC count, neutrophil count and percentage, lymphocyte count, PCT, intensive care unit (ICU) admission rate, 28-day mortality, and 28-day survival (Table 1).

Figure 1 shows the flow chart of study enrolment. Significant between-group differences were observed in the patients’ clinical characteristics (Table 1 and Table 2). The 28-day survival rate was significantly higher in the COVID-19 group than in the non-COVID-19 group (Figure 3).

**Figure 2.**
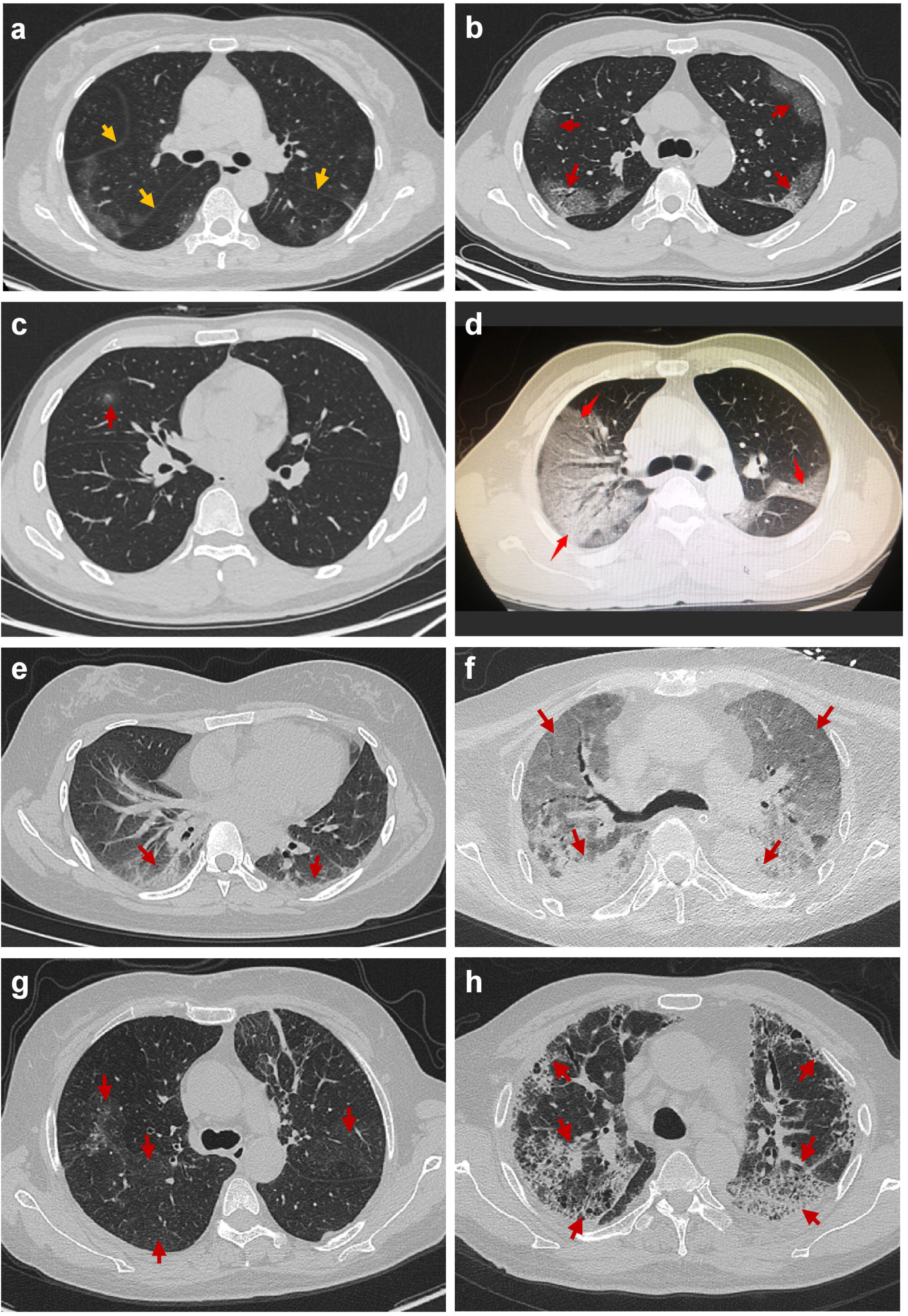

**Figure 3.**
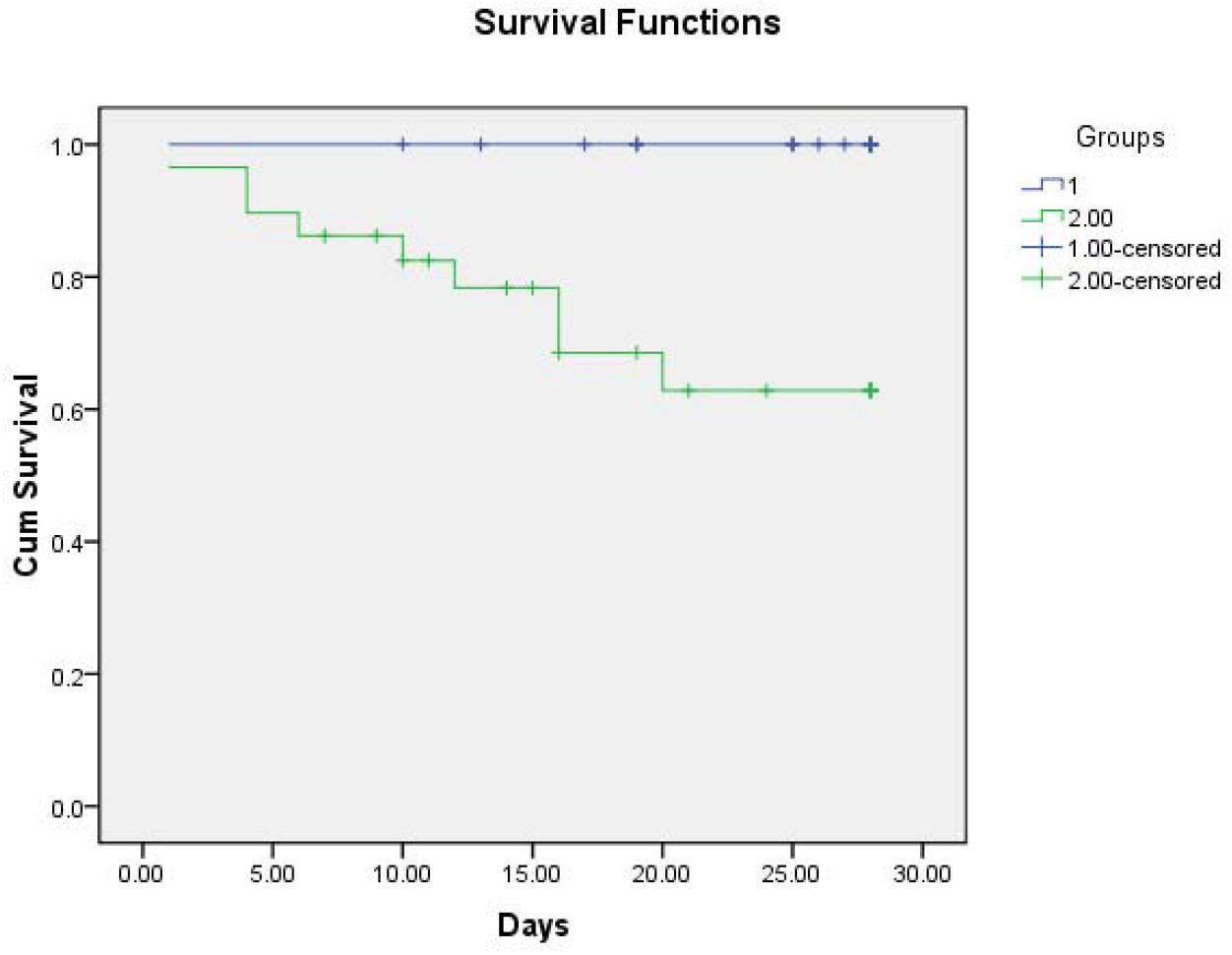
The 28-day survival rates of COVID-19 versus non-COVID-19 patients (higher for COVID-19, P = 0.002)

### Discussion

Within three months of the first COVID-19 patient reported in Wuhan, more than one million confirmed cases have been reported worldwide. Owing to effective prevention and control measures implemented during the first 50 days in China[17], the basic reproduction number (R0) of SARS-CoV-2 has been suppressed from 3.15 on January 23 to 0.04, preventing at least 700,000 cases outside Wuhan. COVID-19 occurred during the peak season for other viral respiratory infections, including viral pneumonia[18]. Therefore, it is important to distinguish COVID-19 from non-COVID-19 cases. In this study, we analysed the clinical data of patients with viral pneumonia who were treated at our hospital during a single period and found that all 23 COVID-19 cases were imported cases. At the initial visit, only 69.57% of the patients had fever, and approximately one-third had no fever. Nucleic acid test results may be positive before symptoms appear [19], posing a great challenge for the early detection of imported COVID-19 cases. Dry cough is a typical symptom of COVID-19, which should be noted during inquiry to prevent missed diagnoses. Both COVID-19 and non-COVID-19 were more common in middle-aged adults, as the mean age was 45 in the COVID-19 group and 53 in the non-COVID-19 group. In both groups, two-thirds of the patients were men. These data are consistent with those of large clinical studies[12, 20]. In addition, most COVID-19 patients (91.30%) had a history of travelling to or from Wuhan or had contact with individuals from Wuhan [21] whereas non-COVID-19 patients had no clear history of contact. COVID-19 patients were younger with fewer underlying diseases whereas non-COVID-19 patients were older with more underlying diseases (such as cerebrovascular disease, organ transplant, haematological tumour, chronic obstructive pulmonary disease, rheumatic disease, diabetes, and immunodeficiency), more severe conditions, a higher ICU admission rate, and a higher 28-day mortality (Table 1). The 28-day survival rate was higher in the COVID-19 group (Figure 2). COVID-19 was more likely to affect T cell immunity, resulting in weakened T cell/cellular immunity. Most COVID-19 cases were mild and presented typical symptoms (such as dry cough with little phlegm). Only three COVID-19 patients were admitted to the ICU.

It is important to determine how to screen and diagnose COVID-19 early to prevent imported nosocomial infections. This study showed that clinical symptoms and blood tests may provide important clues. No significant between-group differences were observed in fever, blood pressure, heart rate, respiratory rate, platelet count, or kidney function. In addition to dry cough, COVID-19 was characterized by lower WBC count, neutrophil count and percentage, and lymphocyte count and percentage, which, combined with the history of exposure to individuals from Hubei and positive nucleic acid test results, may help diagnose COVID-19. In COVID-19 patients, chest CT showed the following characteristics: 1) in most cases, CT showed typical visceral pleural inflammatory exudate on days five to seven of onset (Figure 2A, day six of onset, ‘inflammation line’, yellow arrow); 2) typical CT findings included multiple hazy, patchy ground glass opacities near the visceral pleura in the middle and lower lobes (Figure 2B, day 10 of onset, red arrow); 3) in mild cases, CT showed small, patchy ground glass opacities without clear boundaries (Figure 2C, day two of onset, red arrow); 4) in severe cases, chest CT showed bilateral multiple large, patchy ground glass opacities with consolidation (Figure 2D, day eight of onset, red arrow). Non-COVID-19 was associated with a different time of onset, disease course, disease severities, and chest CT findings. In rhinovirus infection combined with influenza B pneumonia, CT showed bilateral scattered patchy opacities and parenchymal bands with bronchiectasis (Figure 2E, day 14 of onset, red arrow). Cytomegalovirus infection combined with fungal, bacterial, or pneumocystis pneumonia is usually observed after organ (kidney) transplant; in our cytomegalovirus patients, chest CT showed bilateral scattered ground glass opacities with interstitial consolidation (Figure 2F, day 14 of onset, red arrow). In severe influenza A pneumonia, CT showed bilateral diffuse bronchial thickening, with parenchymal bands and consolidation around the bronchi (Figure 2G, day 8 of onset, red arrow). In severe respiratory syncytial virus pneumonia with rheumatoid arthritis, chest CT showed multiple patchy opacities, parenchymal bands, ground glass opacities, consolidation, reticular markings, and interstitial fibrosis in both lungs (Figure 2H, day 10 of onset, red arrow). These characteristics help distinguish COVID-19 from other viral pneumonia infections[22–26].

This study has some limitations. First, this was a retrospective study with a short observation time, so the level of evidence was lower than that of prospective randomized controlled trials. Second, the sample size was small, with only 23 COVID-19 cases, most of which were mild, and only three patients were admitted to the ICU according to our protocol[27]. Non-COVID-19 patients were older with more underlying diseases and thus were more critical, with 12 of these patients admitted to the ICU. The 28-day mortality rate of non-COVID-19 patients was higher than that of COVID-19 patients, and the 28-day survival rate of non-COVID-19 patients was lower than that of COVID-19 patients. Third, no effective treatment is available for COVID-19. We followed the guidelines[28] and administered oxygen therapy, oxygen atomization and humidification, antiviral therapy (lopinavir/ritonavir tablets, Arbidol, interferon nasal spray), ambroxol (to break up phlegm), acetylcysteine (Fluimucil, to prevent pulmonary fibrosis), live combined Bifidobacterium and Lactobacillus tablets (to supplement intestinal probiotics), and α-thymosin for supportive symptomatic treatment in the case of decreased T lymphocytes. Other than traditional Chinese medicine (oral or via a nasal tube), the treatment protocol lacks specifics. Last, prospective studies are necessary to validate our results.

This study shows that a history of contact with individuals from Wuhan, dry cough, low WBC count, low neutrophil count, low lymphocyte count, low T lymphocyte count, typical chest CT findings, and a positive nucleic acid test for SARS-CoV-2 helps screen and diagnose COVID-19, as well as distinguish it from other (non-COVID-19) viral pneumonia infections.

### Conclusion

Epidemiological data, clinical symptoms, nucleic acid test results for COVID-19, and chest CT manifestations may help distinguish COVID-19 from non-COVID-19 cases.

## Data Availability

The data used to support the findings of this study are available from the corresponding author upon request.

## Ethics approval and consent to participate

The clinical research ethics boards of the West China Hospital approved the study, ethics code: 2019(130).

## Authors’ contributions

Xiao-Jin Li conceived and designed the study. Xiao-Jin Li and Bing-Xing Shuai retrieved the medical databases, screened the literature, and extracted the data. Zhong-Wei Zhang conducted quality evaluation and statistical analyses. Yan Kang contributed to validation project administration. The manuscript was drafted and examined by Xiao-Jin Li and Yan Kang.

## Consent for publication

All authors read and approved the final manuscript.

## Acknowledgements

This work was supported by the 1·3·5 Project for disciplines of excellence-clinical Research Incubation Project(2019HXFH061), West China Hospital, Sichuan University, and the Sichuan University Spark Project (2018SCUH0031). The authors thank Professor Xue-Lian Liao, statistician Cai-Rong Zhu and Jing Zhang for their contributions to this study.

